# Economic modelling of providing “spare” adrenaline autoinjectors to all schools to improve the management of anaphylaxis

**DOI:** 10.1101/2025.08.01.25332796

**Authors:** Paul J Turner, Andrew D Bright, Louise J Michaelis, Jennifer K. Quint

## Abstract

**Objective:** To analyse NHS health datasets to estimate the cost of providing emergency adrenaline autoinjectors (AAIs) to school pupils on a named-patient basis to leave on school premises, versus providing “spare” AAIs to schools which can be used in any school pupil.

**Design:** Retrospective cohort study.

**Setting:** English primary electronic health data from the Clinical Practice Research Datalink (CPRD) and English prescriptions data from the NHS Business Services Authority.

**Participants:** School-aged children in England.

**Main outcome measures:** i) Proportion of school children with food allergy prescribed AAI; ii) Cost of providing more than 2 AAIs to individual pupils mapped to Integrated care boards (ICBs) in England, compared to the cost of providing 4 “spare” AAIs to every school, for the academic year 2023/24.

**Results:** 44% of school-aged children in CRPD had at least one AAI prescription, and only 34% had repeat AAIs prescribed. In pupils with previous anaphylaxis, rates were 59% and 44% respectively. During the academic year 2023/24, 63% of pupils were dispensed more than 2 AAIs, at an estimated cost of over £9million. The estimated cost of providing “spare” AAIs to every school was £4.5million. Were “spare” AAIs to replace the supply of named-patient AAIs exclusively to leave on school premises, this would represent a potential cost-saving of at least £4.6million or 25% of the total national expenditure for AAIs

**Conclusions:** Under half of children at risk of anaphylaxis are prescribed AAIs. Providing “spare” AAIs to all schools (at no cost to the school) would be a cost-neutral strategy for the vast majority of ICBs – and one that is likely to improve emergency access to AAIs and therefore safety.

**KEY MESSAGES:** *What is already known on this topic:*

- In 2017, UK legislation was changed to allow schools to obtain, without prescription, “spare” adrenaline autoinjector (AAI) devices for the emergency treatment of anaphylaxis in any school pupil.

*What this study adds:*

- Under half of school-aged children with food allergy (and at potential risk of anaphylaxis) are prescribed AAIs.
- While the MHRA recommends people at risk of anaphylaxis carry 2 AAIs, in school-children prescribed AAIs, over 60% were dispensed more than 4 AAIs in the academic year 2023/24; it is likely that the majority of these additional AAIs were for provided to be left on school premises.
- If “spare” AAIs were provided to all schools, to avoid the need for pupils to leave their own AAIs on school premises, this would represent a potential cost-saving of at least £4.6million or 25% of the total national expenditure for AAIs.

*How this study might affect research, practice or policy:*

- This analysis clearly demonstrates providing “spare” AAIs to schools (at no cost to the school) would be a cost-neutral strategy which would improve emergency access to AAIs for all school pupils (not just the minority prescribed AAIs) and also increase the resilience of the UK supply chain for AAIs.

## INTRODUCTION

Around 3% of school-aged children in the UK have immunoglobulin-E (IgE-)-mediated food allergies.^1^ Thus, on average, UK school classes will have one or two children at risk of food-induced anaphylaxis, a serious allergic reaction which may be life-threatening. Even with the best dietary avoidance, most children will have at least one accidental reaction every 2-3 years.^2–4^ While most will not progress to anaphylaxis, severity is unpredictable which is why people at risk of anaphylaxis are usually prescribed adrenaline autoinjectors (AAI) for emergency use.^5^ The majority of reactions respond to a single dose, but up to 10% require a further dose^6^ and devices may misfire or be used incorrectly. This is why the Medicines and Healthcare Products Regulatory Agency (MHRA) and European Medicines Authority recommend that individuals at risk of food-anaphylaxis have access to two AAIs at all times.^7,8^

School children spend around 20% of their waking hours in school. It is therefore not surprising that 16-18% of school-aged children with food allergies have had a reaction in school.^9,10^ Around 80% of all anaphylaxis reactions to food occur in school-aged children,^11^ and 20% of these happen at school.^12^ One quarter of these anaphylaxis reactions in school occur in pupils with no prior allergy diagnosis.^13^ Fortunately, fatal anaphylaxis is rare,^14^ but it also very unpredictable:^5^ 17% of anaphylaxis deaths in UK school-aged children happen in the educational setting.^14^

To help mitigate this risk, many schools require pupils at risk of anaphylaxis to not only have AAIs with them, but to leave the devices on school premises in case they forget to bring them in. While the MHRA is explicit as to the need to carry two AAIs at all times,^7^ there is less clarity over the number of devices that should be prescribed to school children: the British Society for Allergy & Clinical Immunology (BSACI) are increasingly aware of General Practitioners (GPs) who refuse to prescribe more than 2 devices to any given individual. Many food-allergic children with only prior mild reactions are not prescribed AAI; however, anaphylaxis often happens in those with only previous mild reactions.^5^ Under current UK legislation, an AAI supplied on prescription to any given patient cannot be used in someone else – even in an emergency^15^ – so schools can only use a child’s AAI on that specific child.

In 2015, the BSACI, working with Anaphylaxis UK and Allergy UK, undertook a national survey to evaluate anaphylaxis care in schools. Responses were received from 1609 parents and 821 teachers, with representation from every region across the UK.^16^ Parents reported that 83% of their food-allergic children had been prescribed AAIs to leave on school premises (the majority two devices, although 18% were issued with a single device and 10% were supplied with 3+ devices specifically for school). 93% of teachers worked in a school with at least one child prescribed AAI for school.

It is this background which led to UK law being changed in 2017, to allow for schools to obtain, without a prescription, “spare” AAI devices for use in emergencies (for example where the pupil’s own AAI is not readily available or they do not have their own AAI prescribed).^17^ To support schools, the Department of Health and Social Care (DHSC) together with key stakeholders developed non-statutory guidance.^18^ However, uptake of “spare” AAIs has been limited with only around half of schools doing so^19^ – with schools having to fund the cost of spare AAI directly and pay “market rate” – often in excess of £100 per device (rather than the subsidised NHS tariff, currently £9.90 per 2 devices).

To address this, some Integrated Care Boards (ICBs) have funded local pilots whereby “spare” AAI are provided to local schools.^20^ We analysed NHS health datasets to assess the potential cost of providing “spare” AAIs to schools, and how this might be offset by primary care no longer providing AAIs to individual pupils (on a named-patient basis) to leave on school premises.

## METHODS

### Data Sources and Study Population

Clinical Practice Research Datalink (CPRD) Aurum is a large UK primary care electronic healthcare record database with current data for approximately 20% of the English population; it is considered representative of the English population in terms of age, sex, deprivation, and regional distribution.^21^ We used data from the May 2021 build of CPRD Aurum (including AAI prescription data) and secondary care data from NHS England’s Hospital Episode Statistics (HES) Admitted Patient Care database, to evaluate prescription data for AAI in children ≤18 years between 2008 and 2018.^1^

The NHS Business Services Authority (NHSBSA) is an arm’s length body of the DHSC, delivering a range of national platforms, systems and services to support primary care, the NHS workforce and UK citizens. NHSBSA processes around 1.1 billion NHS prescription items annually, dispensed within a primary care setting. We evaluated NHS prescriptions data relating to AAIs from April 2022 to March 2025. The data were limited to prescribing in primary care in England, which is also dispensed in the community in England. Further information regarding the dataset and caveats over its use can be found in the Supplementary Methods.

### Analyses

We evaluated children/young people aged 5-18 years with a diagnosis of food allergy in CPRD Aurum, as previously described.^1^ Linking this to 2015 Index of Multiple Deprivation (IMD) data for England and secondary health-care data from the HES Admitted Patient Care database, we evaluated potential factors associated with AAI prescription, using a logistic regression model to estimate odds ratios (GraphPad Prism, version 10.4.2).

We then evaluated AAI prescriptions to school pupils of primary (reception–year 6) and secondary school age (year 7–year 11) during the 2023/24 and 2024/25 academic years, using NHSBSA data. Specifically, we assessed the number of pupils prescribed >2 AAIs in the period of interest, in England as a whole and by ICB. Children around 25kg are often switched from a 150mcg to a 300mcg dose: we therefore excluded devices prescribed prior to a change in prescription dose when this was observed within the year of interest. For the most recent academic year 2024/25, data were only available for the 8-months August 2024–March 2025, therefore we estimated annual cost by extrapolating the data to a 12-month period. We assessed the validity of this approach by evaluating monthly dispensing of AAIs, and also applying this method to the academic year 2023/24, where data was available for the full 12-month period.

We estimated the potential annual cost-savings, both overall and by ICB, were ICBs to provide every school in England with four “spare” AAIs on an annual basis (for primary schools, 2 × 150mcg dose and 2 × 300mcg dose in line with DHSC guidance;^18^ for secondary schools, 4 × 300mcg) – rather than supply more than two AAIs to each pupil prescribed AAI in a given year. NHSBSA prescription data do not show why a patient was prescribed an AAI – it could be that they are replacing expired, misplaced or used devices, or provided as additional sets for other settings rather than being supplied as additional AAIs for school use. Given that the in-date period for AAIs is usually at least 12 months, we used a base-assumption that dispensing more than 2 AAIs was for additional supply for school (since used AAI are typically dispensed through hospital pharmacies).^22^ However, we also ran a sensitivity analysis where we assumed that dispensing a single device was more likely to be more replacement while dispensing a pair of devices (2 AAIs) was more likely to be for school, since schools typically request 2 AAIs to be left on the premises per pupil.^16^ This assumption is supported by data showing that 90% of reactions respond to a single dose of adrenaline.^6^ Using this approach, we therefore calculated an estimated minimum and maximum cost-saving. Data relating to the number of schools within each ICB were obtained from the Office for National Statistics.^23^ We assumed the same cost for supplying AAIs to schools as to patients, given that currently, supply to schools of “spare” pens is through community pharmacies.^18^

### Ethics Approval

CPRD has approval for the collection and release of anonymised primary care data for observational research from the NHS Health Research Authority (reference 05/MRE04/87). The protocol for this research was approved by CPRD’s Research Data Governance Process (protocol number: 20_156R2). The analyses conducted by the NHSBSA were undertaken using pseudonymised NHS Prescriptions data within the NHSBSA secure data environment, in accordance with the NHSBSA Privacy Notice and relevant data protection legislation.

## RESULTS

### Frequency of any AAI prescription in school-aged children in CPRD Aurum

28,520 individuals aged 5-18 years (inclusive) had at least one diagnostic code for food allergy in CPRD Aurum and were eligible for linkage with HES data; 21,586 met the definition for probable food allergy.^1^ Overall, 9,567 (44%) had at least one AAI prescribed (Table S1). AAI prescription was more common in children of primary school age (49% of 5-10 year olds) compared with secondary school (40% of 11-18 year olds, p<0.0001, Chi squared). Only 34% (40% of 5-10 years, 28% of 11-18 years; p<0.0001) had a repeat prescription for AAI. Nut allergy and a history of previous anaphylaxis were associated with a higher odds of AAI prescription, whilst increasing age and higher Index of Multiple Deprivation (IMD) were associated with lower odds (Table 1). Being managed exclusively outside the hospital setting was associated with a slightly lower OR for AAI prescription (OR 0.87, p=0.01), but not for repeat prescription.

**Table 1:** Odds ratios [95% confidence interval] for f actors associated with AAI prescription in the Clinical Practice Research Datalink (CPRD) dataset.

| Factor | At least 1 prescription for AAI | Repeat prescription for AAI |
| --- | --- | --- |
| Age | **0.95 [0.94 to 0.95]** | **0.92 [0.91 to 0.93]** |
| Sex | 1.02 [0.96 to 1.08] | 1.00 [0.95 to 1.07] |
| IMD | **0.95 [0.93 to 0.96]** | **0.93 [0.91 to 0.95]** |
| Nut allergy | **6.14 [5.64 to 6.69]** | **6.04 [5.48 to 6.67]** |
| History of previous anaphylaxis | **4.09 [3.50 to 4.81]** | **3.47 [2.96 to 4.06]** |
| Managed exclusively in primary care | 0.87 [0.78 to 0.97] | 0.96 [0.85 to 1.08] |

IMD, Index of Multiple Deprivation. Odds ratio in **bold**, $p < 0.0001$

### Frequency of prescription of more than 2 AAIs

Using NHSBSA prescription data, we found that during the 2023/24 academic year, 63% of school-aged children prescribed AAI were dispensed with 3+ devices, and 60% received 4+ devices. The proportion of AAI prescription items across the whole population that could not be linked to individual patients was 1.3% between August 2023 and March 2025. The estimated cost of providing more than 2 AAIs per person was >£9million, representing almost half of the total ICB expenditure for AAIs in that year (Table 2). Prescription of >2 AAIs was more common for primary school-aged children versus those in secondary school (p<0.0001, chi-squared). Similar patterns were seen for the 8-month period from Aug 2024–Mar 2025. We did not find any significant impact of IMD on rates of dispensing >2 AAIs (data not shown).

**Table 2:**
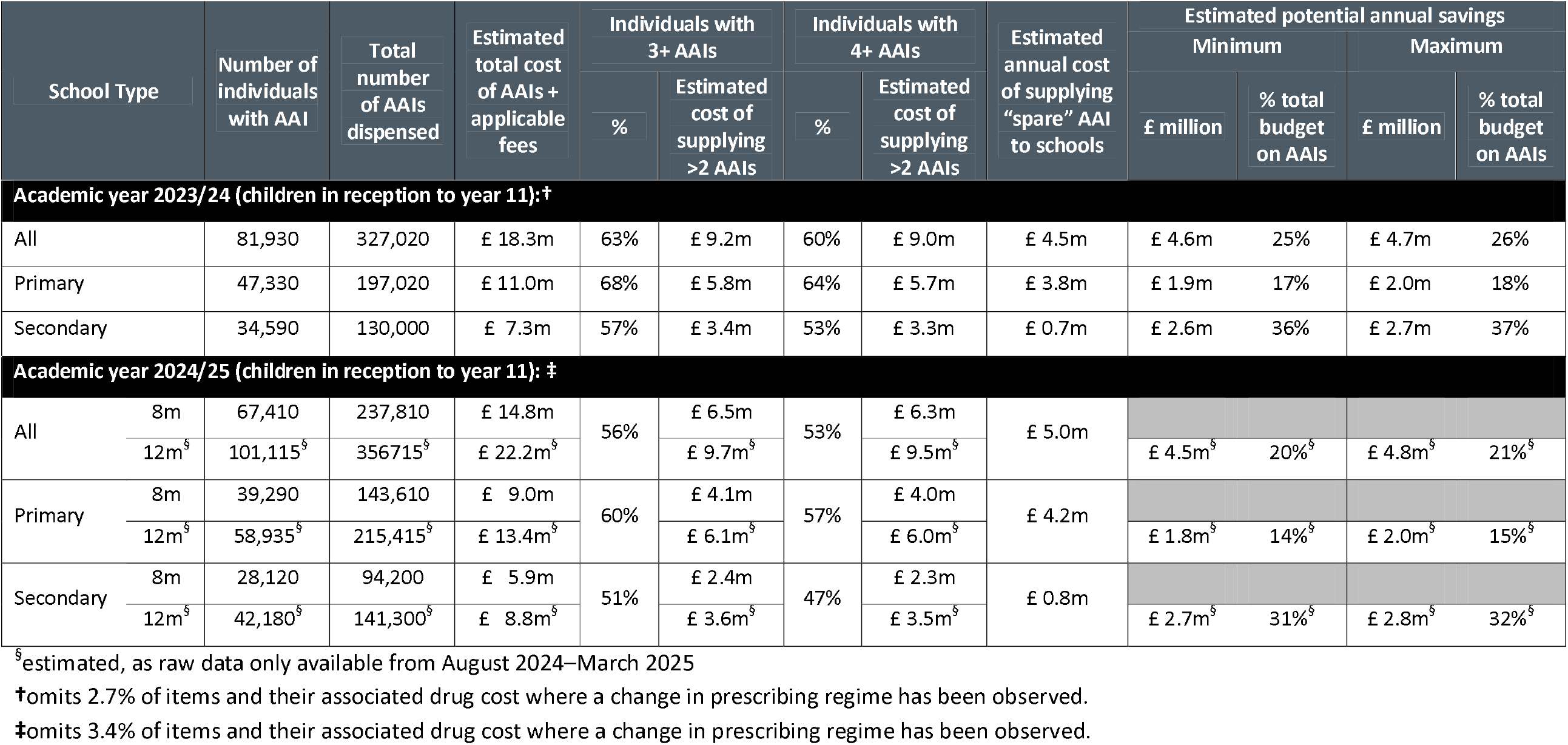
Economic cost of providing “spare” adrenaline autoinjectors to all schools in England, if this cost was offset by the cost of no longer routinely dispensing more than 2 devices to school-aged children.

### Monthly trends in AAI prescribing

At the time of analysis, data for the 2024/25 academic year were available for the first eight months only, as the academic year was still in progress. To provide an annual estimate for the cost of providing “spare” AAI to schools for 2024/25, we assessed if it was reasonable to extrapolate data for August 2024–March 2025 to the entire academic year. To test the validity of this approach, we examined if the number of AAIs dispensed each month was consistent across the year from April 2022 to March 2025. We found evidence for a monthly spike in AAI prescriptions dispensed in September (Figure 1), coinciding with the start of the UK academic year; 13% of all AAI prescriptions were issued in September, instead of an expected monthly average of 8.3%. Given this, we also evaluated the impact of extrapolating 8-months data (August 2023–March 2024) to the entire academic year (August 2023–July 2024), and compare this to the actual data available for the 12 months August 2023–July 2024. This analysis is shown in Table S2. Despite the “September spike”, extrapolating data from August 2023–March 2024 to the full 12-month period provided a reasonable estimate of the number of AAI prescribed and thus the total cost, but under-estimated the proportion of pupils dispensed more than 2 AAIs.

**Figure 1:**
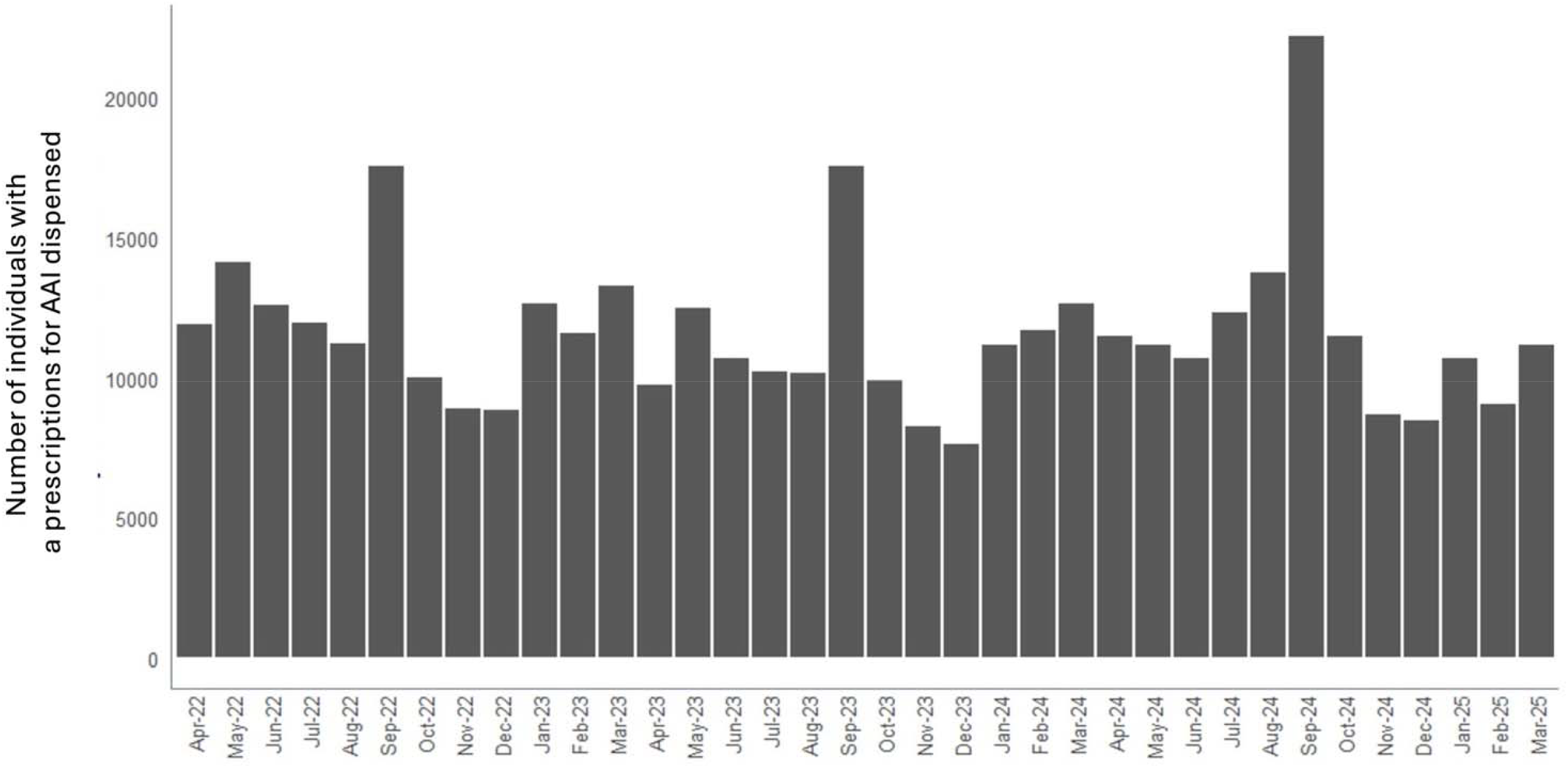
Number of individuals with a prescription for AAI dispensed, by calendar month.

### Economic modelling of providing “spare” adrenaline autoinjectors

Finally, we estimated the relative cost/saving of providing every school in England with four “spare” AAIs on an annual basis, and whether this could be offset by the current cost of dispensing additional AAIs to pupils beyond the two recommended by the MHRA. For the 2023/24 academic year, the cost of providing “spare” AAI to every school was estimated to be £4.5million; the cost of providing more than 2 AAIs on a named-patient basis was >£9million, therefore this represented a potential cost-saving of at least £4.6million or 25% of the total national expenditure for AAIs (Table 2). Estimated savings by ICB are shown in Table S3. Across the 42 ICBs, only 4 (10%) would incur additional significant cost (≥£10,000, approximately 5% of total expenditure on AAIs) while 31 (74%) would achieve cost-savings in excess of £10,000 (and some in excess of £400,000). The average cost-saving per ICB would be over £70,000 (Table S1 and Figure 2). A similar level of savings was also noted for the academic year 2024/25 (Table 2), with estimated savings by individual ICB shown in Table S4.

**Figure 2:**
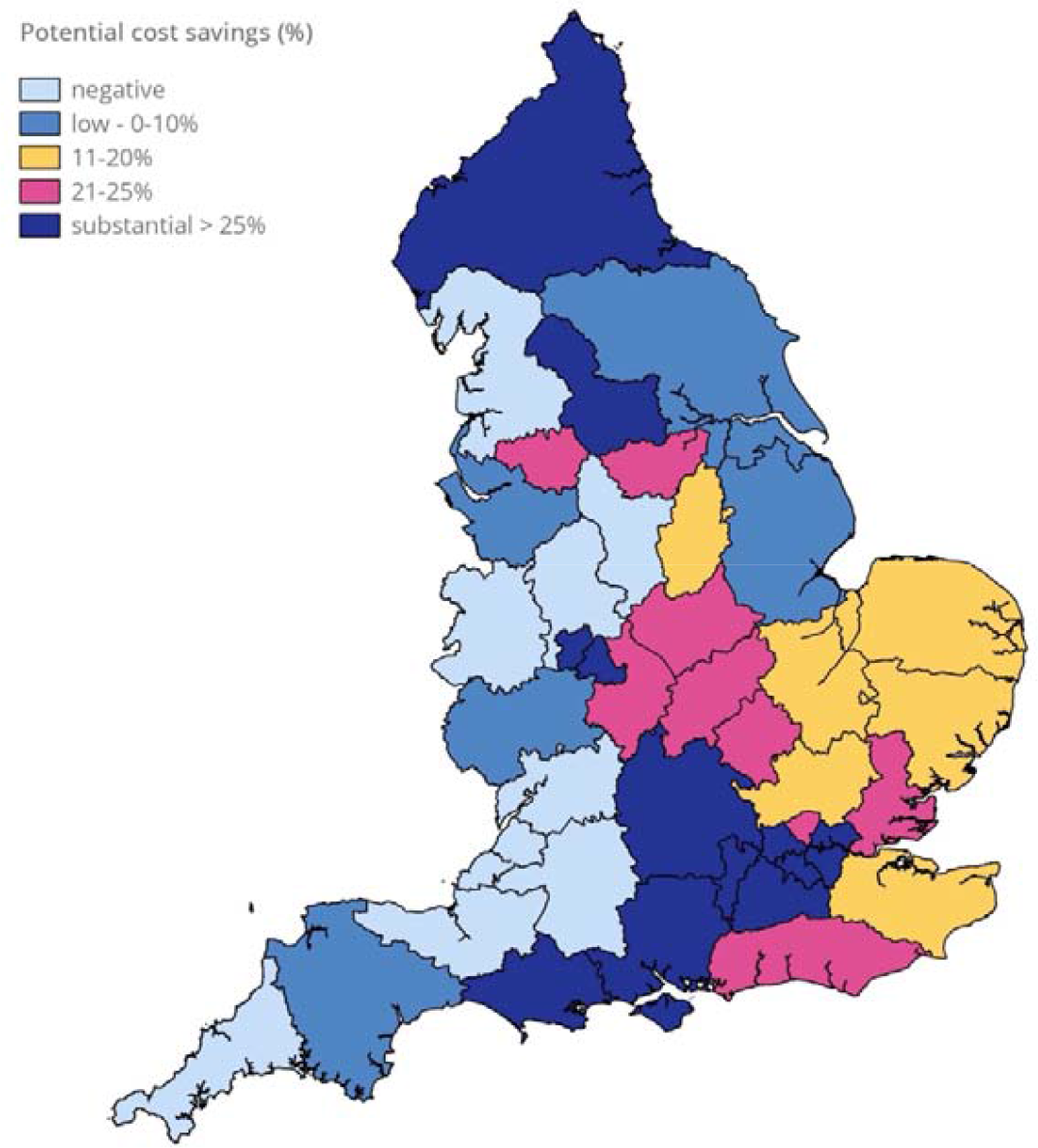
Estimated cost-savings by ICB by limiting general supply of adrenaline auto-injectors on a named patient basis to 2 devices per person, and supplying all schools within the ICB with 4 “spare” devices for emergency use. Data for the academic year 2023/24, as shown in Table S2.

## DISCUSSION

In this analysis of national datasets in England, almost two thirds of school-aged pupils who are prescribed AAI are dispensed with more than >2 devices per annum, with a higher rate in primary school children. Whilst we could not determine the reason for these additional devices, most pupils were dispensed at least another two devices (rather than just a single device). Given that 90% of anaphylaxis reactions respond to a single AAI dose^6^ and that the rate of accidental reactions in nut-allergic children is around 10-15% per annum,^2–4^ this suggests that the majority of additional AAIs dispensed were likely to be additional devices for leaving in school, rather than to replace used AAI.

Since 2017, UK schools have been able to purchase, without a prescription, “spare” AAI devices for emergency use to treat anaphylaxis.^17^ These “spare” devices could be used in any pupil irrespective of whether they had been prescribed AAI, so long as they had an Individualized Health Care Plan (IHCP) and parental consent. Subsequently, the MHRA clarified that “spare” AAIs could be used in any individual (including adults and visitors) in an emergency, but this should be “for exceptional circumstances only that could not have been foreseen”.^24^ However, uptake of “spare” AAIs has been limited,^19^ with the need for schools to pay for the spare AAI themselves as a major factor.^19^ This is in contrast to other schemes in Australia,^25^ Canada^26^ and USA^27^ where “spare” AAIs have been funded centrally.

The need for further change has been flagged by two of His Majesty’s Coroners following inquests into the deaths of Mohammad Ismaeel Ashraf^28^ and Karanbir Cheema^29^ in school, as a result of anaphylaxis. Key concerns highlighted included inadequate staff training resulting in delayed and incorrect administration of adrenaline, and a failure to ensure AAI were in-date and accessible in an emergency – issues which can be addressed through mandatory provision of spare AAIs and training.^30^

For the vast majority of ICBs, we estimated that the cost of spare AAIs to all schools could be fully offset by the ICB no longer funding AAIs on a named-patient basis exclusively for school use – something entirely consistent with both current legislation^17^ and guidance from the MHRA.^31^ There are additional reasons to support such a strategy. Less than 10% of food-allergic children are seen in a specialist allergy clinic;^1^ therefore, many children may not have their risk of anaphylaxis assessed by someone with the requisite experience. In a 2012 survey of 2439 school nurses in the USA, 25.3% of food-allergic students had no AAI and only 24.6% had two unexpired devices at school.^32^ In a survey of 5683 US schools in 2013/14, 607 (11%) reported 919 anaphylaxis events; 22% happened in pupils with no known allergies. 54 pupils (9%) received a second AAI dose.^13^ The NSW Anaphylaxis Education Program in Australia was established in 2004, to improve state-wide anaphylaxis care following several deaths due to anaphylaxis in schools. This included providing “spare” AAIs to all schools.^33^ Between 2017–2019, 341 students had anaphylaxis, of whom 130 (38%) were treated with a “spare” AAI.^34^ Reasons for using the “spare” AAI included: AAI prescribed but not with the child in school or expired (n=17, 5% of anaphylaxis events); no known prior allergies (77, 23%); or known diagnosis of food allergy but AAI not prescribed (36, 11%). By providing all schools with “spare” AAI, all school pupils will be able to access potentially life-saving adrenaline in an emergency.

A further benefit, noted by the DHSC, is that providing “spare” AAI to schools allows schools to hold just a single brand of AAI and avoids the school having to have multiple devices produced by different manufacturers; this reduces confusion over how to use the device (given that instructions differ between brands).^18^ In an emergency, staff can waste valuable minutes identifying a child’s own AAIs, since they cannot use those belonging to someone else. The presence of different brands of AAIs can be confusing, leading to delays in administration as flagged in some inquests.^28,29^ Providing “spare” AAI reduces the time wasted in trying to identify a given child’s own AAI, in an emergency situation where minutes can matter and delays in treatment are associated with fatal outcomes.^30^

Our analysis is not without limitations: there are a number of caveats regarding the use of NHSBSA data. Primarily, the dataset only includes primary care NHS prescriptions in England and dispensed by community pharmacies, and excludes AAI dispensed through hospitals and private healthcare. We were not able to extend this analysis to Scotland, Wales, and Northern Ireland. We could not analyse the reason for dispensing more than two AAIs to any given individual: therefore, we cannot determine whether additional AAIs were dispensed to replace expired/lost devices or to be used in other settings, or to supply additional devices for school use. Notwithstanding, given the spike in the number of AAIs dispensed at the beginning of the school year, and that most of these are for 2+ devices (rather than singlets), it is likely that the majority of “additional” AAIs were for school use rather than to replace used devices.

Schools currently obtain “spare” AAI by placing a request through local pharmacies.^18^ We could therefore assume that the cost of providing “spare” AAI to schools was equivalent to those dispensed on a per-patient basis. Were ICBs to provide “spare” AAI, this might occur through an alternative distribution arrangement, which could affect cost. However, there may be significant advantages: centralised supply may allow schools to be issued AAI from the same batch, meaning the devices would have the same expiry date. This would reduce the burden on schools and allow for more systematic replacement. Centralised distribution would also facilitate monitoring of allergic reactions in school, and help learning from incidents (something already required by UK legislation): such a system has been critical to the success of the NSW Anaphylaxis Education Program in Australia, improving the care of allergic students in schools.^33,34^ Mandatory education of school staff is an essential part of the scheme – an ongoing issue in the UK, which has also been repeatedly flagged as a concern.^19,20,28,29^

Irrespective, there can be little doubt that were ICBs to limit dispensing to two unexpired AAIs per pupil at any one time (and so no longer provide additional AAIs on a named-patient basis just for school use), then providing “spare” AAIs to schools (at no cost to the school) would be a cost-neutral strategy for the vast majority of ICBs – and one that is likely to improve emergency access to AAIs and therefore safety. This would also increase the resilience of the UK supply chain for AAIs (something which has been a major concern in the past decade, and a contributory factor in at least one fatality)^35^ and reduce wastage. While over 2.3 million AAI devices are sold each year, only ~2% are actually used.^7^

In 2020, an editorial concluded that providing spare AAIs to schools can “be achieved with minimal cost implications: with mandatory “spare” AAI provision, families would no longer need to provide the school with a supply of AAIs for each child, something which would avoid confusion and delay in an anaphylaxis emergency… It is what children with food allergy and their families deserve.” ^30^ Five years later, how many more children need to die in UK schools before this is implemented in the UK?^36^

## ACKNOWEDGEMENTS

We thank Lucy Sherwin-Robson, Thomas Owen and colleagues at the NHS Business Service Authority (NHSBSA) who provided NHS Prescriptions data and advice regarding its interpretation. We are also grateful to Constantinos Kallis and Eimear O’Rourke for their assistance in extracting data from CPRD.

## Contributors

PJT, ADB and LJM conceived and designed the study. JKQ acquired the data from CPRD, and conducted initial analyses of that data which were then reviewed by PJT. NHSBSA data was acquired by LSR and TO (contributors), and summary data analysed by LSR, TO and PJT. All authors interpreted the data and reviewed the final version. The guarantor (PJT) accepts full responsibility for the work and the conduct of the study, had access to the summary data and controlled the decision to publish. The corresponding author (PJT) attests that all listed authors meet authorship criteria and that no others meeting the criteria have been omitted.

## Funding

This research was funded by the UK Medical Research Council (reference MR/W018616/1) and the UK Food Standards Agency (reference FS101222). The funders had no role in considering the study design or in the collection, analysis, interpretation of data, writing of the report, or decision to submit the article for publication.

## Competing interests

All authors have completed the ICMJE uniform disclosure form at www.icmje.org/coi_disclosure.pdf and declare: grants from UK Medical Research Council and UK Food Standards Agency for the submitted work; PJT reports grants from Natasha Allergy Research Foundation, JM Charitable Foundation and NIHR/Imperial Biomedical Research Centre, outside the submitted work; personal fees from UpToDate, Elsevier, Stallergenes, outside the submitted work; he is vice-chair of the National Allergy Strategy Group and co-lead of the UK Resuscitation Council Anaphylaxis Working Group. LJM reports Commercial Clinical Trial Research Grants and Advisory Board fees from Danone Nutricia and Regeneron, outside the submitted work. JKQ has received institutional grants from the MRC, NIHR, Health Data Research, GlaxoSmithKline (GSK), BI, Astra Zeneca (AZ), Insmed and Sanofi; and personal fees from GSK, Evidera, Chiesi, AZ and Insmed; and consulting fees from GSK, BI, Sanofi, Chiesi and AZ, outside the submitted work. ADB reports no conflicts of interest.

## Ethics approval

CPRD has approval for the collection and release of anonymised primary care data for observational research from the NHS Health Research Authority (reference 05/MRE04/87). The protocol for this analysis was approved by CPRD’s Research Data Governance Process (protocol number: 20_156R2). The analyses conducted by the NHSBSA were undertaken in accordance with the NHSBSA Privacy Notice and relevant data protection legislation.

## Data availability statement

Data may be obtained from a third party and are not publicly available. Data may be obtained from a third party and are not publicly available. Data sets used in this analysis were obtained via a Clinical Practice Research Datalink (CPRD) institutional licence. Requests for data should be made directly to the CPRD via their online application portal (https://cprd.com/research-applications).

---

**Table S1:** Provision of AAI in children aged 5-18 years in CPRD Aurum.

|  | <u>**Overall cohort**</u> | <u>**5-10 years**</u> | <u>**11-18 years**</u> |
| --- | --- | --- | --- |
| **Probable food allergy** | **21,586** | **10,721** | **10,865** |
| • At least 1 AAI prescribed | 9567 (44%) | 5204 (49%) | 4363 (40%) |
| • Repeat prescription for AAI | 7300 (34%) | 4262 (40%) | 3038 (28%) |
| **Nut allergy specified** | **14,277** | **6910** | **7367** |
| • At least 1 AAI prescribed | 7441 (52%) | 3993 (58%) | 3448 (47%) |
| • Repeat prescription for AAI | 5727 (40%) | 3292 (48%) | 2435 (33%) |
| **Previous anaphylaxis specified** | **996** | **326** | **670** |
| • At least 1 AAI prescribed | 584 (59%) | 220 (68%) | 364 (54%) |
| • Repeat prescription for AAI | 436 (44%) | 191 (59%) | 245 (37%) |

**Table S2:**
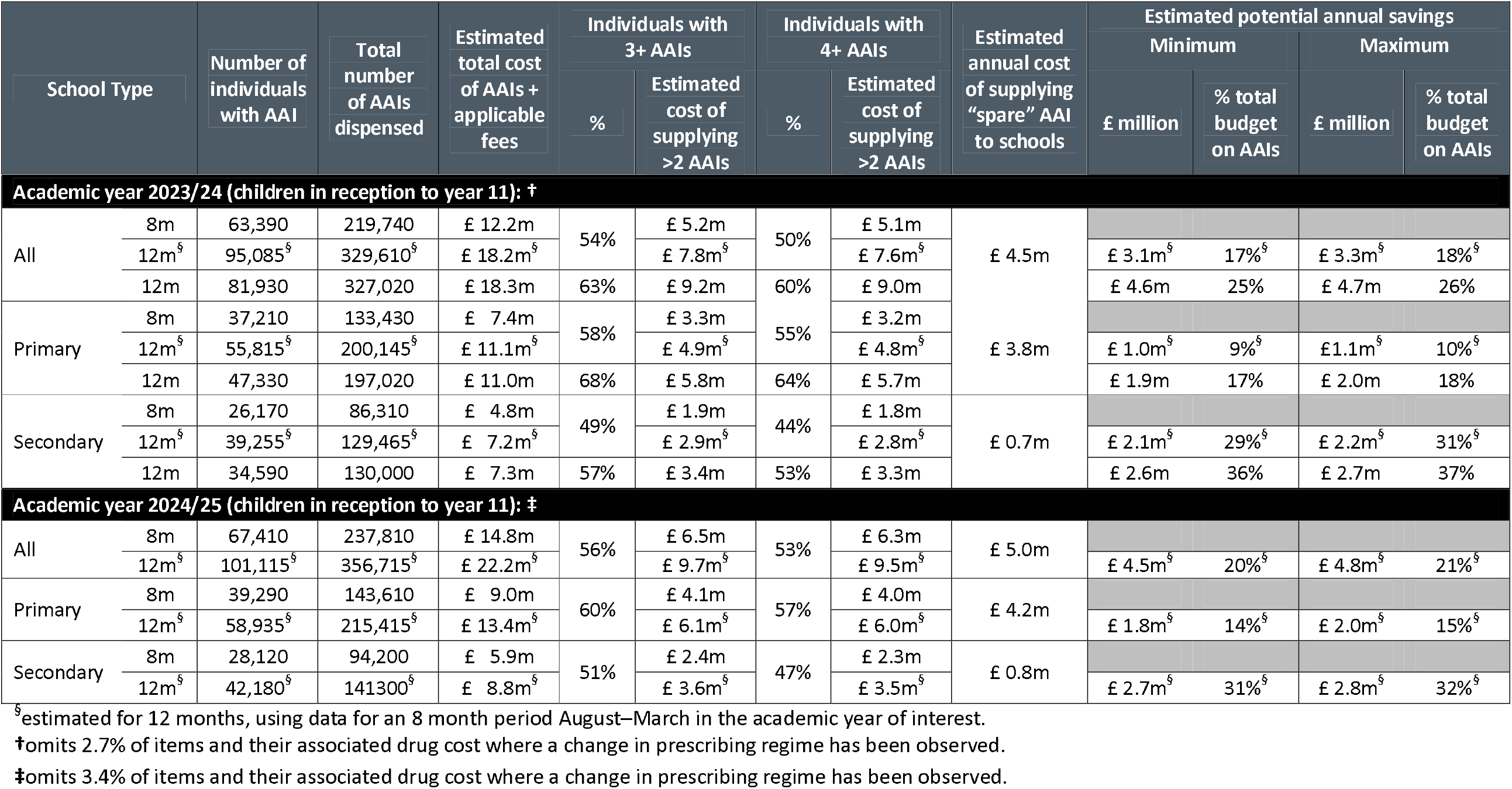
Sensitivity analysis evaluating the impact of using 8 months of data (August-March) to estimate potential annual savings for the 12 months between August-July in each academic year of interest.

**Table S3:** Economic cost of providing “spare” adrenaline autoinjectors to all schools in England, if this cost was offset by the cost of no longer routinely dispensing more than 2 devices to school-aged children, for the Academic year 2023/24, by ICB.^* †^.

| ICB | Number of individuals with AAI | Total number of AAls dispensed | Estimated total cost of AAls + applicable fees | Individuals with 3+ AAls |  | Individuals with 4+ AAls |  | Number of schools in ICB | Estimated annual cost of supply for "spare" AAI to schools | Estimated potential annual savings |  |  |  |
| --- | --- | --- | --- | --- | --- | --- | --- | --- | --- | --- | --- | --- | --- |
|  |  |  |  | % | Estimated cost of supplying >2 AAls | % | Estimated cost of supplying >2 AAls |  |  | Minimum |  | Maximum |  |
|  |  |  |  |  |  |  |  |  |  | £ | % total budget on AAls | £ | % total budget on AAls |
| NHS Bath and NE Somerset, Swindon and Wiltshire | 830 | 3,050 | £ 170,1k | 59% | £ 79,3k | 54% | £ 77,1k | 390 | £ 87,3k | (£ 10,2k) | -6% | (£ 7,9k) | -5% |
| NHS Bedfordshire, Luton and Milton Keynes | 1,720 | 6,120 | £ 342,1k | 57% | £ 151,7k | 52% | £ 147,3k | 340 | £ 76,1k | £ 71,2k | 21% | £ 75,7k | 22% |
| NHS Birmingham and Solihull | 2,970 | 11,200 | £ 627,4k | 56% | £ 297,4k | 49% | £ 286,7k | 449 | £ 100,5k | £ 186,3k | 30% | £ 196,9k | 31% |
| NHS Black Country | 1,760 | 6,840 | £ 382,8k | 59% | £ 189,0k | 55% | £ 184,5k | 406 | £ 90,8k | £ 93,7k | 24% | £ 98,1k | 26% |
| NHS Bristol, N Somerset and S Gloucestershire | 890 | 3,040 | £ 170,8k | 54% | £ 72,6k | 48% | £ 69,8k | 307 | £ 68,7k | £ 1,1k | 1% | £ 3,9k | 2% |
| NHS Buckinghamshire, Oxfordshire and Berkshire West | 3,630 | 14,390 | £ 803,1k | 65% | £ 400,9k | 60% | £ 391,9k | 684 | £ 153,1k | £ 238,9k | 30% | £ 247,8k | 31% |
| NHS Cambridgeshire and Peterborough | 1,220 | 4,490 | £ 250,0k | 59% | £ 116,9k | 54% | £ 113,5k | 323 | £ 72,3k | £ 41,2k | 16% | £ 44,6k | 18% |
| NHS Cheshire and Merseyside | 2,100 | 8,490 | £ 475,7k | 65% | £ 242,8k | 57% | £ 233,8k | 900 | £ 201,4k | £ 32,4k | 7% | £ 41,4k | 9% |
| NHS Cornwall & Isles of Scilly | 570 | 2,130 | £ 118,9k | 60% | £ 56,3k | 58% | £ 55,7k | 267 | £ 59,7k | (£ 4,0k) | -3% | (£ 3,5k) | -3% |
| NHS Coventry and Warwickshire | 1,360 | 5,250 | £ 293,8k | 63% | £ 142,4k | 58% | £ 139,0k | 337 | £ 75,4k | £ 63,6k | 22% | £ 67,0k | 23% |
| NHS Derby and Derbyshire | 930 | 3,650 | £ 204,1k | 65% | £ 100,3k | 62% | £ 99,1k | 487 | £ 109,0k | (£ 9,8k) | -5% | (£ 8,7k) | -4% |
| NHS Devon | 1,260 | 4,650 | £ 260,2k | 60% | £ 120,8k | 56% | £ 118,6k | 474 | £ 106,1k | £ 12,5k | 5% | £ 14,8k | 6% |
| NHS Dorset | 920 | 3,710 | £ 206,7k | 71% | £ 104,6k | 68% | £ 103,5k | 224 | £ 50,1k | £ 53,4k | 26% | (£ 54,5k) | 26% |
| NHS Frimley | 1,700 | 6,030 | £ 337,8k | 55% | £ 149,7k | 51% | £ 145,8k | 239 | £ 53,5k | £ 92,3k | 27% | £ 96,2k | 28% |
| NHS Gloucestershire | 500 | 1,770 | £ 98,69k | 56% | £ 43,5k | 52% | £ 42,3k | 285 | £ 63,8k | (£ 21,4k) | -22% | (£ 20,3k) | -21% |
| NHS Greater Manchester | 2,670 | 11,840 | £ 663,4k | 66% | £ 366,3k | 63% | £ 361,3k | 1,023 | £ 228,9k | £ 132,4k | 20% | £ 137,4k | 21% |
| NHS Hampshire and I. of Wight | 2,650 | 11,510 | £ 642,5k | 74% | £ 347,3k | 72% | £ 344,5k | 600 | £ 134,3k | £ 210,2k | 33% | £ 213,0k | 33% |
| NHS Herefordshire and Worcs | 690 | 2,760 | £ 154,1k | 67% | £ 78,1k | 61% | £ 75,8k | 299 | £ 66,9k | £ 8,9k | 6% | £ 11,2k | 7% |
| NHS Hertfordshire & W. Essex | 3,420 | 11,410 | £ 639,7k | 47% | £ 258,8k | 45% | £ 254,3k | 591 | £ 132,2k | £ 122,0k | 19% | £ 126,5k | 20% |
| NHS Humber and N. Yorkshire | 1,740 | 6,860 | £ 383,0k | 64% | £ 190,3 | 60% | £ 186,3k | 719 | £ 160,9k | £ 25,5k | 7% | £ 29,4k | 8% |
| NHS Kent and Medway | 2,450 | 9,170 | £ 512,7k | 60% | £ 240,3k | 56% | £ 235,3k | 654 | £ 146,3k | £ 89,0k | 17% | £ 94,0k | 18% |
| NHS Lancashire & S. Cumbria | 1,210 | 4,740 | £ 265,2k | 60% | £ 131,2k | 57% | £ 129,0k | 773 | £ 173,0k | (£ 44,0k) | -17% | (£ 41,8k) | -16% |
| NHS Leicester, Leics and Rutland | 1,850 | 6,620 | £ 370,2k | 55% | £ 165,5k | 48% | £ 157,6k | 395 | £ 88,4k | £ 69,2k | 19% | £ 77,1k | 21% |
| NHS Lincolnshire | 660 | 2,670 | £ 149,4k | 64% | £ 76,1k | 59% | £ 74,4k | 333 | £ 74,5k | (£ 0,1k) | 0% | £ 1,6k | 1% |
| NHS Mid and South Essex | 2,210 | 7,690 | £ 430,0k | 52% | £ 185,0k | 49% | £ 181,0k | 381 | £ 85,3k | £ 95,8k | 22% | £ 99,7k | 23% |
| NHS Norfolk and Waveney | 1,060 | 4,410 | £ 245,1k | 69% | £ 128,9k | 65% | £ 126,6k | 440 | £ 98,5k | £ 28,2k | 11% | £ 30,4k | 12% |
| NHS North Central London | 3,620 | 15,220 | £ 853,3k | 65% | £ 448,8k | 63% | £ 444,3k | 1,208 | £ 270,3k | £ 174,0k | 20% | £ 178,5k | 21% |
| NHS NE London | 4,330 | 18,080 | £1,012,9k | 62% | £ 530,6k | 59% | £ 524,5k | 372 | £ 83,2k | £ 441,2k | 44% | £ 447,4k | 44% |
| NHS NE and N Cumbria | 3,280 | 14,270 | £ 797,4k | 69% | £ 433,6k | 66% | £ 428,5k | 503 | £ 112,6k | £ 316,0k | 40% | £ 321,0k | 40% |
| NHS NW London | 4,970 | 19,970 | £1,118,1k | 65% | £ 565,8k | 61% | £ 552,4k | 491 | £ 109,9k | £ 442,5k | 40% | £ 456,0k | 41% |
| NHS Northamptonshire | 1,040 | 4,210 | £ 233,6k | 67% | £ 119,0k | 63% | £ 116,8k | 297 | £ 66,5k | £ 50,3k | 22% | £ 52,5k | 22% |
| NHS Nottingham and Notts | 1,270 | 5,100 | £ 285,8k | 63% | £ 143,4k | 60% | £ 141,1k | 420 | £ 94,0k | £ 47,1k | 16% | £ 49,4k | 17% |
| NHS Shropshire, Telford & Wrekin | 380 | 1,580 | £ 88,1k | 66% | £ 45,8k | 66% | £ 45,8k | 213 | £ 47,7k | (£ 1,9k) | -2% | (£ 1,9k) | -2% |
| NHS Somerset | 410 | 1,420 | £ 79,5k | 49% | £ 33,8k | 49% | £ 33,8k | 246 | £ 55,0k | (£ 21,3k) | -27% | (£ 21,3k) | -27% |
| NHS SE London | 4,350 | 19,200 | £1,074,5k | 72% | £ 589,3k | 70% | £ 584,3k | 486 | £ 108,7k | £ 475,5k | 44% | £ 480,5k | 45% |
| NHS SW London | 3,790 | 15,080 | £ 844,5k | 65% | £ 421,8k | 61% | £ 414,5k | 390 | £ 87,3k | £ 327,2k | 39% | £ 334,5k | 40% |
| NHS S Yorkshire | 1,400 | 5,890 | £ 330,6k | 65% | £ 175,3k | 64% | £ 174,1k | 477 | £ 106,7k | £ 67,4k | 20% | £ 68,5k | 21% |
| NHS Staffordshire and Stoke-on-Trent | 930 | 3,570 | £ 199,4k | 57% | £ 96,8k | 54% | £ 95,1k | 444 | £ 99,3k | (£ 4,2k) | -2% | (£ 2,6k) | -1% |
| NHS Suffolk and NE Essex | 1,280 | 4,800 | £ 267,3k | 59% | £ 125,9k | 55% | £ 122,6k | 375 | £ 83,9k | £ 38,7k | 14% | £ 42,0k | 16% |
| NHS Surrey Heartlands | 2,580 | 10,810 | £ 604,1k | 73% | £ 316,2k | 70% | £ 311,7k | 305 | £ 68,2k | £ 243,5k | 40% | £ 247,9k | 41% |
| NHS Sussex | 2,200 | 9,140 | £ 510,5k | 69% | £ 267,2k | 65% | £ 262,7k | 497 | £ 111,2k | £ 151,5k | 30% | £ 156,0k | 31% |
| NHS West Yorkshire | 3,190 | 14,190 | £ 793,7k | 71% | £ 439,0k | 67% | £ 432,9k | 875 | £ 195,8k | £ 237,1k | 30% | £ 243,2k | 31% |

\*omits 3.4% of items and their associated drug cost where a change in prescribing regime has been observed.
†omits <0.01% of patients whose practice could not be aligned to an ICS.

**Table S4:** Economic cost of providing “spare” adrenaline autoinjectors to all schools in England, if this cost was offset by the cost of no longer routinely dispensing more than 2 devices to school-aged children, from August – March during the Academic year 2024/25, by ICB.^* †^.

| ICB | Number of individuals with AAI | Total number of AAls dispensed | Estimated total cost of AAls + applicable fees | Individuals with 3+ AAls |  | Individuals with 4+ AAls |  | Number of schools in ICB | Estimated annual cost of supply for “spare” AAI to schools | Estimated potential annual savings |  |  |  |
| --- | --- | --- | --- | --- | --- | --- | --- | --- | --- | --- | --- | --- | --- |
|  |  |  |  | % | Estimated cost of supplying >2 AAls | % | Estimated cost of supplying >2 AAls |  |  | Minimum |  | Maximum |  |
|  |  |  |  |  |  |  |  |  |  | £ | % total budget on AAls | £ | % total budget on AAls |
| NHS Bath and NE Somerset, Swindon and Wiltshire | 720 | 2,410 | £ 140,0k | 56% | £ 61,3k | 49% | £ 58,2k | 390 | £ 97,3k | (£ 10,0k) | -7% | (£ 5,3k) | -4% |
| NHS Bedfordshire, Luton and Milton Keynes | 1,420 | 4,520 | £ 281,7k | 50% | £ 107,3k | 45% | £ 103,0k | 340 | £ 84,8k | £ 69,7k | 25% | £ 76,2k | 27% |
| NHS Birmingham and Solihull | 2,580 | 8,460 | £ 528,0k | 48% | £ 209,2k | 41% | £ 197,3k | 449 | £ 112,0k | £ 184,0k | 35% | £ 201,8k | 38% |
| NHS Black Country | 1,550 | 5,470 | £ 340,8k | 53% | £ 149,0k | 47% | £ 143,4k | 406 | £ 101,2k | £ 113,8k | 33% | £ 122,2k | 36% |
| NHS Bristol, N Somerset and S Gloucestershire | 740 | 2,300 | £ 143,2k | 45% | £ 51,5k | 41% | £ 49,6k | 307 | £ 76,6k | (£ 2,1k) | -1% | £ 0,7k | 0% |
| NHS Buckinghamshire, Oxfordshire and Berkshire West | 2,870 | 9,970 | £ 621,5k | 57% | £ 264,9k | 52% | £ 256,2k | 684 | £ 170,6k | £ 213,7k | 34% | £ 226,8k | 36% |
| NHS Cambridgeshire and Peterborough | 920 | 3,070 | £ 190,8k | 52% | £ 78,0k | 48% | £ 75,5k | 323 | £ 80,5k | £ 32,6k | 17% | £ 36,4k | 19% |
| NHS Cheshire and Merseyside | 1,700 | 5,940 | £ 370,4k | 58% | £ 60,7k | 51% | £ 153,8k | 900 | £ 224,4k | £ 6,3k | 2% | £ 16,6k | 4% |
| NHS Cornwall & Isles of Scilly | 440 | 1,510 | £ 93,7k | 55% | £ 39,2k | 50% | £ 37,9k | 267 | £ 66,6k | (£ 9,7k) | -10% | (£ 7,8k) | -8% |
| NHS Coventry and Warwickshire | 1,160 | 3,850 | £ 240,0k | 53% | £ 97,1k | 49% | £ 94,0k | 337 | £ 84,0k | £ 57,0k | 24% | £ 61,6k | 26% |
| NHS Derby and Derbyshire | 740 | 2,430 | £ 151,0k | 51% | £ 60,0k | 49% | £ 58,7k | 487 | £ 121,4k | (£ 33,4k) | -22% | (£ 31,5k) | -21% |
| NHS Devon | 1,090 | 3,750 | £ 233,7k | 57% | £ 99,8k | 54% | £ 97,9k | 474 | £ 118,2k | £ 28,7k | 12% | £ 31,5k | 13% |
| NHS Dorset | 740 | 2,780 | £ 173,0k | 68% | £ 81,5k | 65% | £ 80,3k | 224 | £ 55,9k | £ 64,6k | 37% | £ 66,4k | 38% |
| NHS Frimley | 1,350 | 4,240 | £ 264,5k | 45% | £ 98,2k | 40% | £ 93,9k | 239 | £ 59,6k | £ 81,2k | 31% | £ 87,8k | 33% |
| NHS Gloucestershire | 400 | 1,300 | £ 80,6k | 53% | £ 31,5k | 48% | £ 30,3k | 285 | £ 71,1k | (£ 25,6k) | -32% | (£ 23,8k) | -29% |
| NHS Greater Manchester | 2,140 | 8,040 | £ 501,2k | 58% | £ 237,2k | 54% | £ 231,6k | 1,023 | £ 255,1k | £ 92,3k | 18% | £100,750 | 20% |
| NHS Hampshire and I. of Wight | 2,170 | 8,440 | £ 525,2k | 68% | £ 256,5k | 66% | £ 254,0k | 600 | £ 149,6k | £ 231,4k | 44% | £235,149 | 45% |
| NHS Herefordshire and Worcs | 540 | 1,910 | £ 118,7k | 59% | £ 51,3k | 54% | £ 49,4k | 299 | £ 74,6k | £ 0,4k | 0% | £2,391 | 2% |
| NHS Hertfordshire & W. Essex | 2,760 | 8,130 | £ 507,4k | 39% | £ 165,3k | 36% | £ 159,7k | 591 | £ 147,4k | £ 92,1k | 18% | £100,548 | 20% |
| NHS Humber and N. Yorkshire | 1,410 | 4,920 | £ 305,9k | 56% | £ 131,2k | 53% | £ 128,7k | 719 | £ 179,3k | £ 13,8k | 5% | £17,510 | 6% |
| NHS Kent and Medway | 2,010 | 6,830 | £ 425,3k | 54% | £ 177,3k | 50% | £ 173,0k | 654 | £ 163,1k | £ 96,4k | 23% | £ 102,9k | 24% |
| NHS Lancashire & S. Cumbria | 980 | 3,520 | £ 219,4k | 53% | £ 98,0k | 51% | £ 96,7k | 773 | £ 192,8k | (£ 47,7k) | -22% | (£ 45,8k) | -21% |
| NHS Leicester, Leics and Rutland | 1,430 | 4,490 | £ 280,1k | 47% | £ 104,3k | 38% | £ 96,8k | 395 | £ 98,5k | £ 46,7k | 17% | £ 57,9k | 21% |
| NHS Lincolnshire | 520 | 1,770 | £ 110,1k | 52% | £ 46,7k | 48% | £ 45,4k | 333 | £ 83,0k | (£ 14,9k) | -14% | (£ 13,0k) | -12% |
| NHS Mid and South Essex | 1,830 | 5,620 | £ 350,2k | 44% | £ 125,9k | 40% | £ 121,6k | 381 | £ 95,0k | £ 87,3k | 25% | £93,889 | 27% |
| NHS Norfolk and Waveney | 780 | 2,720 | £ 169,1k | 56% | £ 73,3k | 55% | £ 72,7k | 440 | £ 109,7k | £ 0,7k) | 0% | £ 0,2k | 0% |
| NHS North Central London | 3,100 | 11,510 | £ 718,2k | 60% | £ 332,2k | 58% | £ 327,2k | 1,208 | £ 301,2k | £ 189,6k | 26% | £ 197,1k | 27% |
| NHS NE London | 3,880 | 14,860 | £ 927,2k | 60% | £ 445,5k | 57% | £ 437,4k | 372 | £ 92,8k | £ 563,3k | 61% | £ 575,4k | 62% |
| NHS NE and N Cumbria | 2,470 | 9,020 | £ 562,5k | 59% | £ 255,9k | 56% | £ 251,5k | 503 | £ 125,4k | £ 251,8k | 45% | £ 258,3k | 46% |
| NHS NW London | 4,150 | 14,600 | £ 910,7k | 58% | £ 398,7k | 54% | £ 387,4k | 491 | £ 122,4k | £ 458,7k | 50% | £ 475,5k | 52% |
| NHS Northamptonshire | 840 | 2,840 | £ 176,7k | 54% | £ 73,2k | 49% | £ 70,7k | 297 | £ 74,1k | £ 32,1k | 18% | £ 35,8k | 20% |
| NHS Nottingham and Notts | 1,050 | 3,770 | £ 235,2k | 56% | £ 104,7k | 54% | £ 103,4k | 420 | £ 104,7k | £ 50,4k | 21% | £ 52,3k | 22% |
| NHS Shropshire, Telford & Wrekin | 310 | 1,050 | £ 65,2k | 55% | £ 27,5k | 55% | £ 27,5k | 213 | £ 53,1k | (£ 11,8k) | -18% | (£ 11,8k) | -18% |
| NHS Somerset | 380 | 1,160 | £ 71,9k | 45% | £ 25,7k | 39% | £ 24,5k | 246 | £ 61,3k | (£ 24,6k) | -34% | (£ 22,8k) | -32% |
| NHS SE London | 3,670 | 14,450 | £ 901,3k | 68% | £ 445,3k | 66% | £ 441,0k | 486 | £ 121,2k | £ 540,3k | 60% | £ 546,8k | 61% |
| NHS SW London | 3,160 | 11,300 | £ 705,4k | 59% | £ 313,9k | 56% | £ 308,3k | 390 | £ 97,3k | £ 365,2k | 52% | £ 373,6k | 53% |
| NHS S Yorkshire | 1,180 | 4,410 | £275,4k | 58% | £ 128,5k | 55% | £ 126,0k | 477 | £ 118,9k | £ 70,1k | 25% | £ 73,8k | 27% |
| NHS Staffordshire and Stoke-on-Trent | 780 | 2,560 | £ 159,1k | 49% | £ 63,4k | 45% | £ 61,6k | 444 | £ 110,7k | (£ 18,4k) | -12% | (£ 15k,6) | -10% |
| NHS Suffolk and NE Essex | 1,010 | 3,280 | £ 203,4k | 50% | £ 80,4k | 46% | £ 77,9k | 375 | £ 93,5k | £ 23,4k | 12% | £ 27,2k | 13% |
| NHS Surrey Heartlands | 2,010 | 7,390 | £460,2k | 64% | £ 210,6k | 61% | £ 207,5k | 305 | £ 76,1k | £ 235,1k | 51% | £ 239,8k | 52% |
| NHS Sussex | 1,750 | 6,330 | £ 393,9k | 59% | £ 177,4k | 56% | £ 173,6k | 497 | £ 123,9k | £ 136,5k | 35% | £ 142,1k | 36% |
| NHS West Yorkshire | 2,740 | 10,930 | £ 681,2k | 66% | £ 342,2k | 63% | £ 337,2k | 875 | £ 218,2k | £ 287,6k | 42% | £ 295,1k | 43% |

\* omits 2.7% of items and their associated drug cost where a change in prescribing regime has been observed.
† omits <0.01% of patients whose practice could not be aligned to an ICS.

## Supplementary Methods

The analyses of the estimated number of school age children receiving AAIs and associated quantities and costs are based on electronic and paper-based NHS prescriptions dispensed in the community in England. This does not include medicines used in secondary care, prisons, issued by a private prescriber, or over-the-counter. The dataset was used to estimate the number of school-aged children in school years Reception to Year 11, who received AAI prescriptions (coded as per Table S1) during the academic years 2023/24 and 2024/25 (incomplete, data only available to March 2025). The dataset also quantifies the cost of the prescriptions, as well as the estimated quantity and associated costs for patients receiving more than two AAIs. The dataset is aggregated at a national level, ICB level and by patient IMD.

**Table S1:**
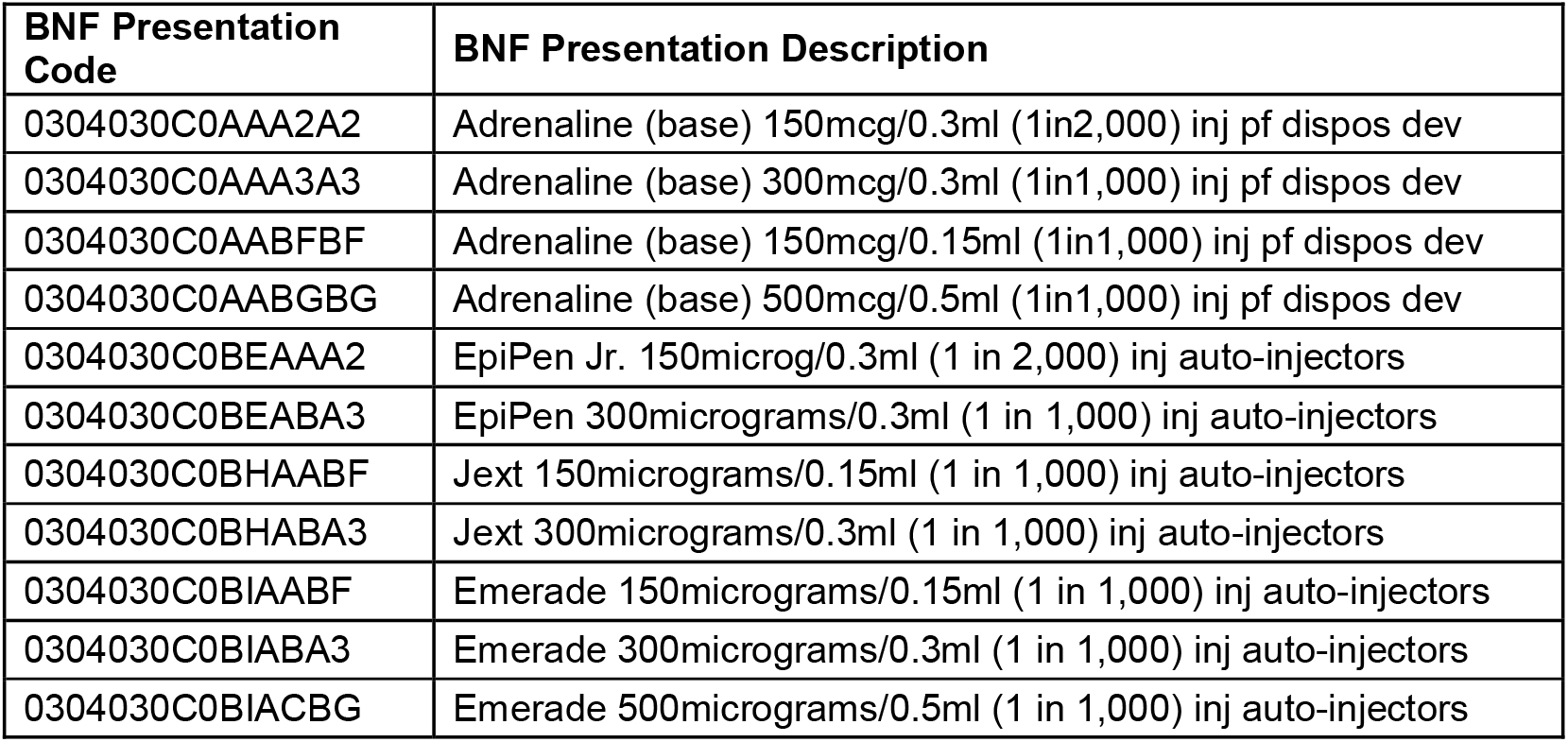
BNF Prescription codes used in this analysis.

| BNF Presentation Code | BNF Presentation Description |
| --- | --- |
| 0304030C0AAA2A2 | Adrenaline (base) 150mcg/0.3ml (1in2,000) inj pf dispos dev |
| 0304030C0AAA3A3 | Adrenaline (base) 300mcg/0.3ml (1in1,000) inj pf dispos dev |
| 0304030C0AABFBF | Adrenaline (base) 150mcg/0.15ml (1in1,000) inj pf dispos dev |
| 0304030C0AABGBG | Adrenaline (base) 500mcg/0.5ml (1in1,000) inj pf dispos dev |
| 0304030C0BEAAA2 | EpiPen Jr. 150microg/0.3ml (1 in 2,000) inj auto-injectors |
| 0304030C0BEABA3 | EpiPen 300micrograms/0.3ml (1 in 1,000) inj auto-injectors |
| 0304030C0BHAABF | Jext 150micrograms/0.15ml (1 in 1,000) inj auto-injectors |
| 0304030C0BHABA3 | Jext 300micrograms/0.3ml (1 in 1,000) inj auto-injectors |
| 0304030C0BIAABF | Emerade 150micrograms/0.15ml (1 in 1,000) inj auto-injectors |
| 0304030C0BIABA3 | Emerade 300micrograms/0.3ml (1 in 1,000) inj auto-injectors |
| 0304030C0BIACBG | Emerade 500micrograms/0.5ml (1 in 1,000) inj auto-injectors |

## Data caveats

The following caveats apply to all NHSBSA data:

- NHSBSA data cannot confirm why a patient received a prescription nor what the remaining shelf life of their medication was when they received their prescription. NHSBSA prescriptions data can show the number of school aged children who have received an AAI dispensed in the community, the number of AAIs received and the cost of these AAIs. NHSBSA data does not show why the AAI were prescribed to a patient, for example, whether they are replacing expired pens, used pens, misplaced pens or whether they are receiving additional pens to be kept in school. It also does not show instances where pens were dispensed in the hospital.
- Data are limited to prescriptions where an NHS number has been captured during NHSBSA processing. The proportion of AAI prescription items across the whole population that could not be linked to individual patients was 1.3% between August 2023 and March 2025. This means that data used in the analysis represents most, but not all, patients.
- Data are limited to prescribing in primary care in England which is dispensed in the community in England.
- Prescriptions excluded from the data are:
  ∘ Items not dispensed, disallowed and those returned to the contractor for further clarification.
  ∘ Prescriptions issued and dispensed in Prisons, Hospitals and through private practice.
  ∘ Items prescribed but not presented for dispensing or not submitted to NHS Prescription Services by the dispenser.

The following caveats apply to the data used in the analysis:

- When a child reaches the recommended switching age, their prescribed dose may increase from 150mcg to 300mcg/500mcg. In such cases, only the updated stronger doses are considered for analysis. Conversely, if a child receives a 300mcg dose followed by a 150mcg dose, we still count only the devices linked to the most recent dose. This approach ensures we do not count any devices dispensed prior to any changes in a patient’s prescribing regime.
- Data are limited to AAI prescriptions which were submitted to the NHSBSA for payment between August 2023 and March 2025. Prescribing intervals are: 12 months (Aug 2023– Jul 2024) and 8 months (Aug 2024–Mar 2025). The month in NHSBSA data relates to the dispensing month for which the prescription batch was submitted. This is generally, but not always, the month in which the prescription was dispensed. This means that there may be dispensing that has not been submitted to the NHSBSA for payment and is therefore not included.
- For the Academic year 2024/25, the 8-month time interval captures children between Reception and Year 11. Patients were only included if their date of birth was between 1^st^ September 2008 and 31^st^ August 2020. The 12-month time interval during the 2023/24 academic year captured children between Reception and Year 11 only. Patients are only included in these analyses if their date of birth was between 1^st^ September 2007 and 31^st^ August 2019.
  ∘ As these are estimated quantities, figures are rounded up or down to the nearest 10 throughout to reflect a degree of uncertainty.
  ∘ School type reporting is based on the school year a child would have been in during the 23/24 or 24/25 academic year (depending on the chosen time interval), with Reception – Year 6 grouped together as Primary and Year 7 to Year 11 grouped as Secondary.
- ICB reporting is based on the ICB of the practice from which a patient received most of their AAI devices. If a patient’s practice is not captured during processing, they are omitted from the ICB reporting analysis.
- Deprivation reporting is based on the patient’s residential address supplied from their latest auto adrenaline injector prescription. Where address data wasn’t captured directly from the prescription data, we have assigned the address based on other available data for the patient from other prescriptions and Personal Demographic Service (PDS). If no residential address information is available for a patient, they are omitted from the deprivation analysis.

Analysis was limited to the following metrics:

- Number of individuals with AAI: The estimated number of patients based on the count of the unique pseudonymised patient IDs who were dispensed an AAI in the given time period.
- Total number of AAIs dispensed: The sum of AAI devices received by patients.
- Estimated Total Cost of AAIs plus Applicable Fees: The sum of the Net Ingredient Cost (NIC, this relates solely to the basic price of the devices, in the quantity prescribed on a prescription form) plus the applicable prescription charge fees. Applicable fees included are professional fees and consumable container allowance.
- Estimated Cost of Supplying More than 2 AAIs: Determined by summing the total NIC plus fees cost, for all less two AAIs prescribed to a patient. The amount calculated is based on the patient’s average AAI cost, multiplied by the number of AAI devices they received less 2.
- Estimated Proportion of Patients with 3+ AAIs: The estimated number of patients, based on the number of unique pseudonymised patient IDs, where the patient received 3 or more devices divided by the total number of unique pseudonymised patient IDs who received any quantity of AAIs, then multiplied by 100.
- Estimated Proportion of Patients with 4+ AAIs: The estimated number of patients, based on the number of unique pseudonymised patient IDs, where the patient received 4 or more devices divided by the total number of unique pseudonymised patient IDs who received any quantity of AAIs, then multiplied by 100.

## Notes

### Author Declarations

CPRD has approval for the collection and release of anonymised primary care data for observational research from the NHS Health Research Authority (reference 05/MRE04/87). The protocol for this analysis was approved by CPRD Research Data Governance Process (protocol number: 20_156R2). The analyses conducted by the NHSBSA were undertaken in accordance with the NHSBSA Privacy Notice and relevant data protection legislation.

